# Hydroxychloroquine in the treatment of outpatients with mildly symptomatic COVID-19: A multi-center observational study

**DOI:** 10.1101/2020.08.20.20178772

**Authors:** Andrew Ip, Jaeil Ahn, Yizhao Zhou, Andre H. Goy, Eric Hansen, Andrew L Pecora, Brittany A Sinclaire, Urszula Bednarz, Michael Marafelias, Shivam Mathura, Ihor S Sawczuk, Joseph P. Underwood, David M. Walker, Rajiv Prasad, Robert L. Sweeney, Marie G. Ponce, Samuel La Capra, Frank J. Cunningham, Arthur G. Calise, Bradley L. Pulver, Dominic Ruocco, Greggory E. Mojares, Michael P. Eagan, Kristy L. Ziontz, Paul Mastrokyriakos, Stuart L Goldberg

**Author notes:** **Corresponding Author:** Stuart L Goldberg MD, John Theurer Cancer Center at Hackensack University Medical Center, 92 Second Street, Hackensack NJ 07601.

## Abstract

**Background:** Hydroxychloroquine has not been associated with improved survival among hospitalized COVID-19 patients in the majority of observational studies and similarly was not identified as an effective prophylaxis following exposure in a prospective randomized trial. We aimed to explore the role of hydroxychloroquine therapy in mildly symptomatic patients diagnosed in the outpatient setting.

**Methods:** We examined the association between outpatient hydroxychloroquine exposure and the subsequent progression of disease among mildly symptomatic non-hospitalized patients with documented SARS-CoV-2 infection. The primary outcome assessed was requirement of hospitalization. Data was obtained from a retrospective review of electronic health records within a New Jersey USA multi-hospital network. We compared outcomes in patients who received hydroxychloroquine with those who did not applying a multivariable logistic model with propensity matching.

**Results:** Among 1274 outpatients with documented SARS-CoV-2 infection 7.6% were prescribed hydroxychloroquine. In a 1067 patient propensity matched cohort, 21.6% with outpatient exposure to hydroxychloroquine were hospitalized, and 31.4% without exposure were hospitalized. In the primary multivariable logistic regression analysis with propensity matching there was an association between exposure to hydroxychloroquine and a decreased rate of hospitalization from COVID-19 (OR 0.53; 95% CI, 0.29, 0.95). Sensitivity analyses revealed similar associations. QTc prolongation events occurred in 2% of patients prescribed hydroxychloroquine with no reported arrhythmia events among those with data available.

**Conclusions:** In this retrospective observational study of SARS-CoV-2 infected non-hospitalized patients hydroxychloroquine exposure was associated with a decreased rate of subsequent hospitalization. Additional exploration of hydroxychloroquine in this mildly symptomatic outpatient population is warranted.

**Lay Summary:** In this observational study of 1,274 COVID-19 patients, hydroxychloroquine given as an outpatient treatment was associated with a 47% reduction in the hazard of hospitalization. Adverse events were not increased (2% QTc prolongation events, 0% arrhythmias). Further validation is required. Use of hydroxychloroquine to treat COVID-19 in the outpatient setting should be reserved for a clinical trial or after discussion with a physician regarding risks and benefits.

## Introduction

The majority of infections with SARS-CoV-2 result in mildly symptomatic or asymptomatic illnesses that can be managed in outpatient settings. However, progression of the COVID-19 illness may result in significant morbidity and mortality requiring hospitalization and consumption of healthcare resources. In New Jersey, an early COVID-19 epicenter in the United States, approximately 11% of positive cases required hospitalization (216 per 100,000 population).^1^ As testing availability has increased and testing practices have broadened to include mildly symptomatic and asymptomatic individuals the Centers for Disease Control and Prevention has reported a United States national cumulative COVID-19 hospitalization rate of 94.5 per 100,000 individuals.^2^

Hydroxychloroquine, an antimalarial agent with antiviral and anti-inflammatory properties, has been touted as a potential therapy for COVID-19.^3^ Among hospitalized COVID-19 patients, observational studies have noted that hydroxychloroquine exposure has not been associated with a reduction in the risk of death. ‘ A recent observational study from Michigan, however, reported improved survival when hydroxychloroquine was administered within 2 days of hospitalization.^8^ When used as post-exposure prophylaxis within 4 days after moderate or high risk exposure, a prospective randomized trial found that hydroxychloroquine failed to prevent illness compatible with Covid-19 or confirmed infection.^9^

Given that the majority of SARS-CoV-2 infected patients are mildly symptomatic and are managed in the outpatient setting, it remains important to explore whether early administration of hydroxychloroquine could delay progression to more severe illness requiring hospitalization. A trial from Spain randomized younger (mean age 41.6 years) mildly symptomatic outpatients to a 7-day course of hydroxychloroquine or observation, reporting no significant reductions in mean viral load or reduction in hospitalization rate (7.1% control versus 5.9% hydroxychloroquine).^10^ A second randomized study enrolled 491 USA and Canadian subjects via the internet, of whom 34% had virology confirmed infection. Although the overall hospitalization rate was only 3.2% within the population participating in the study (median age 40), more patients receiving placebo (4.7%) compared to hydroxychloroquine (1.9%) required hospitalization.^11^ A Brazilian study of 636 symptomatic, but virology unconfirmed patients treated by telemedicine at home, also noted a reduction in hospitalization rate (5.4% vs 1.9%), with the greatest reductions occurring among the patients who started hydroxychloroquine therapy within the first 7 days of symptoms.^12^ A small French report noted a reduction in symptoms with early therapy compared to observation.^13^ Finally, a German report of 141 outpatients, when compared to cases in the community, noted a decrease in hospitalization rate (2.8% vs 15.4%) with a combination of hydroxychloroquine, azithromycin and zinc.^14^ In summary, the majority of studies, although underpowered to show differences, are all directionally in favor of a reduced hospitalization rate with early outpatient treatment.

Understanding the limitations of observational studies, but with the urgency for evaluating potential therapeutic approaches during the current COVID-19 pandemic, our hospital spanning New Jersey USA established an observational database utilizing an integrated electronic health record (EHR) system (EPIC; Verona, WI).^15-18^ In this multi-center observational cohort study we report progression from mildly symptomatic SARS-CoV-2 infection diagnosed as an outpatient progressing to subsequent need for in-patient hospitalization according to outpatient exposure to hydroxychloroquine.

## Methods

### Study Design and Cohort Selection

This retrospective, observational, multicenter cohort study within the Hackensack Meridian Health network (HMH) utilized EHR-derived data of patients with documented SARS-CoV-2 infection who received care initially within an outpatient setting. Our primary objective was to evaluate the association between hydroxychloroquine exposure and subsequent need for hospitalization in a population of patients with documented SARS-CoV-2 infection diagnosed in the outpatient setting.

Database inclusion and exclusion criteria for this review: 1) Positive SARS-CoV-2 diagnosis by reverse-transcriptase polymerase chain reaction, 2) Outpatient status (includes emergency room diagnosis without immediate hospitalization on the same day) at an HMH outpatient facility between March 1, 2020 until April 22, 2020. Follow-up continued through May 22, 2020.

Institutional Review Board (IRB) approval was obtained for access to the prospective observational database. The requirement for patient informed consent was waived by the IRB as this project represented a non-interventional study utilizing routinely collected data for secondary research purposes.

### Data Sources

We collected data from HMH’s EHR (Epic) which is utilized throughout the network. Outpatients treated at a network related facility were flagged by the EHR if SARS-CoV-2 polymerase chain reaction tests were positive. These EHR-generated reports served as our eligible cohort sample. Demographic, clinical characteristics, treatments, and outcomes were manually abstracted by research nurses and physicians from the John Theurer Cancer Center at Hackensack University Medical Center. Assignment of patients to our data team occurred in real-time but was not randomized. To reduce sampling bias the final cohort included 100% of outpatients by April 22, 2020 as noted on the EHR-generated reports. Data abstracted by the team were entered utilizing Research Electronic Data Capture (REDCap). Quality control was performed by physicians (AI, SLG) overseeing nurse or physician abstraction.

Demographic information was collected by an electronic face-sheet. Comorbidities were defined as diagnosed prior to hospitalization for COVID-19. If not listed in the patient’s record comorbidities were recorded as absent.

### Exposure

For hydroxychloroquine, exposure was defined as a prescription written for the drug as found in the EHR, by documentation in a provider note or in the medication section of the chart. No confirmation of prescription fill or adherence to the medication regimen was attempted. If no evidence of administration of the drug was found, this was recorded as not having received the drug. Hydroxychloroquine exposure, for the purpose of this study, was limited to initiation of treatment in the outpatient setting. Patients who did not have a prehospital exposure, subsequently admitted to a hospital, and then received hydroxychloroquine started in the inpatient setting were counted as having no outpatient exposure to hydroxychloroquine.

### Outcome Measures

The primary outcome measurement was subsequent need for hospitalization with follow-up until May 22, 2020. Hospitalization was identified on EHR review which includes the 13-hospitals within the Hackensack Meridian Health network. The EPIC system also notifies a limited number of participating hospitals outside the network (Epic Care-Everywhere). No attempt to contact the patient to confirm hospitalization outside the network was permitted or performed. Among patients who were hospitalized, the time from date of diagnosis to hospitalization and the requirement for intensive unit care level support or death was also collected. Safety events including discontinuation due to QTc prolongation or arrhythmia incidence after hydroxychloroquine exposure were recorded as per chart review.

Exploratory outcomes included the effect of outpatient hydroxychloroquine exposure on elderly patients over age 65, on patients with more than 2 days of self-reported symptoms, and on patients with at least one reported symptom of fever, shortness of breath, or cough.

### Statistical Analysis

Demographic and clinical parameters of hydroxychloroquine treatment were summarized using median (Q1-Q3) for continuous variables and frequency (percentages) for categorical variables. The differences in the median/distributions of demographic and clinical parameters between the hydroxychloroquine treated and untreated (no hydroxychloroquine) groups were compared using Mood’s median test for continuous variables and Fisher’s exact test or Pearson’s chi-squared test for categorical variables. The comparator group in both the unmatched and propensity matched cohorts included only patients who did not receive hydroxychloroquine.

Multivariable adjusted logistic regression models were fitted to estimate the association between hydroxychloroquine exposure and the need for subsequent hospitalization using clinically likely confounders including age, gender, cancer, hypertension, COPD/asthma, diabetes, fever, cough, shortness of breath, and qSOFA score. When the model goodness-of-fit was not satisfied, we further reduced the aforementioned confounders using the stepwise variable selection and the lasso variable selection.^20^ The hazard ratios (OR) and their 95% confidence intervals were summarized.

To reduce the confounding effects secondary to imbalances in receiving hydroxychloroquine treatment inherent to a retrospective cohort study, we employed propensity-score matching. First, we calculated a propensity score (PS) of receiving hydroxychloroquine treatment for each patient using multivariable logistic regression via adjusting for the aforementioned set of confounder variables except time to hydroxychloroquine treatment. Goodness-of-fit of the multivariable logistic model was examined using the Hosmer-Lemeshow test. We then employed a nonparametric nearest neighbor matching of propensity scores to generate a matched cohort in a 1:10 ratio to pair a patient with hydroxychloroquine treatment to ten patients without hydroxychloroquine treatment (MatchIt Package in R)^19, 20^

With the propensity matched cohort, we repeated the adjusted logistic model with the propensity matched set similar to the unmatched analyses. Sensitivity analyses for confounders were conducted by including the propensity score as a covariate in the unmatched model and by including informative confounders chosen by stepwise selection. Missing data in categorical covariates were coded as a missing data category and were included in the all analyses. Completely observed data by excluding patients with missing covariates were also examined summarized in Supplementary Table. The Kaplan-Meier method and log-rank test were performed to evaluate and compare the median time from date of diagnosis to hospitalization between the hydroxychloroquine treated and untreated groups. Furthermore, we performed an exploratory analysis from time of symptom onset to date of first dose of hydroxychloroquine. A cut-off of less than 2 days from time of symptom onset was used for a logistic regression analysis comparing those with early disease versus later as there appeared to be a stronger benefit to early administration of hydroxychloroquine.^21^ Statistical significance was determined when two-sided p-value<0.05. Subgroup analyses were performed exploratory and thus multiple-test correction was not applied. All statistical analyses were conducted using R software (ver. 3.4., R Project for Statistical Computing).

## Results

### Characterization of the study cohort

There were 4302 patients flagged in the EHR with polymerase chain reaction confirmed infection with SARS-CoV-2. 1274 (30%) patients were evaluated and treated in the outpatient setting prior to any COVID-19 related hospitalization. 97 patients (7.6%) received prescriptions for hydroxychloroquine or had notation of an outpatient exposure to hydroxychloroquine. (Figure 1) Given potential imbalances in treatment allocation due to the observational nature of the study a propensity matched sample was constructed consisting of 1067 patients in total (97 with hydroxychloroquine exposure and 970 without). The distribution of baseline characteristics is shown in Table 1. In the unmatched cohort patients exposed to hydroxychloroquine were more likely to have comorbid conditions. The propensity matched cohorts were well balanced except for an excess of cancer history and a trend towards older age in the hydroxychloroquine cohort.

**Figure 1.**
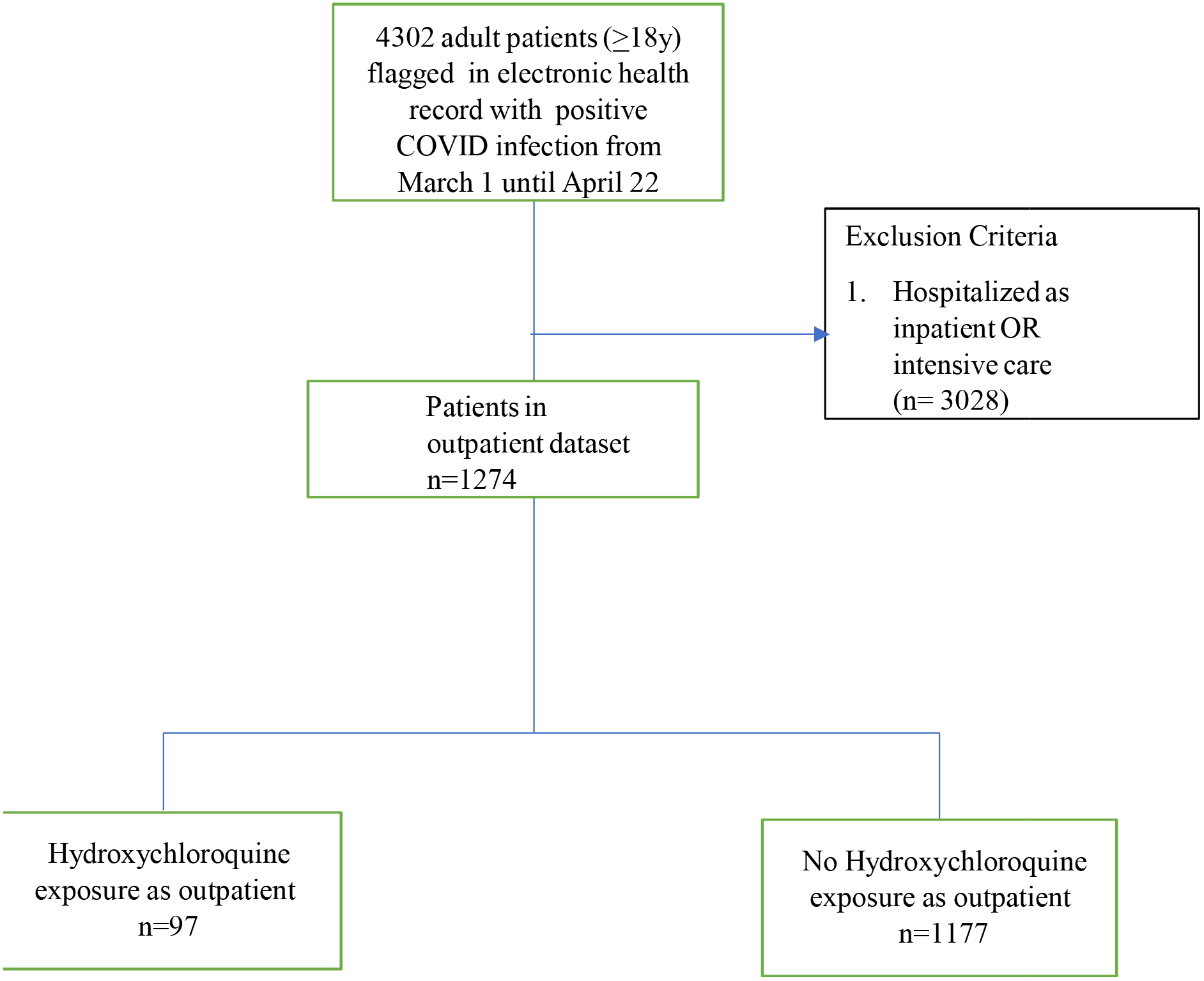
Cohort Selection Flow Diagram. Flow diagram of patient sampling strategy of non-hospitalized COVID-19 patients in Hackensack Meridian Health Network. Follow up occurred until May 22.

**TABLE 1.**
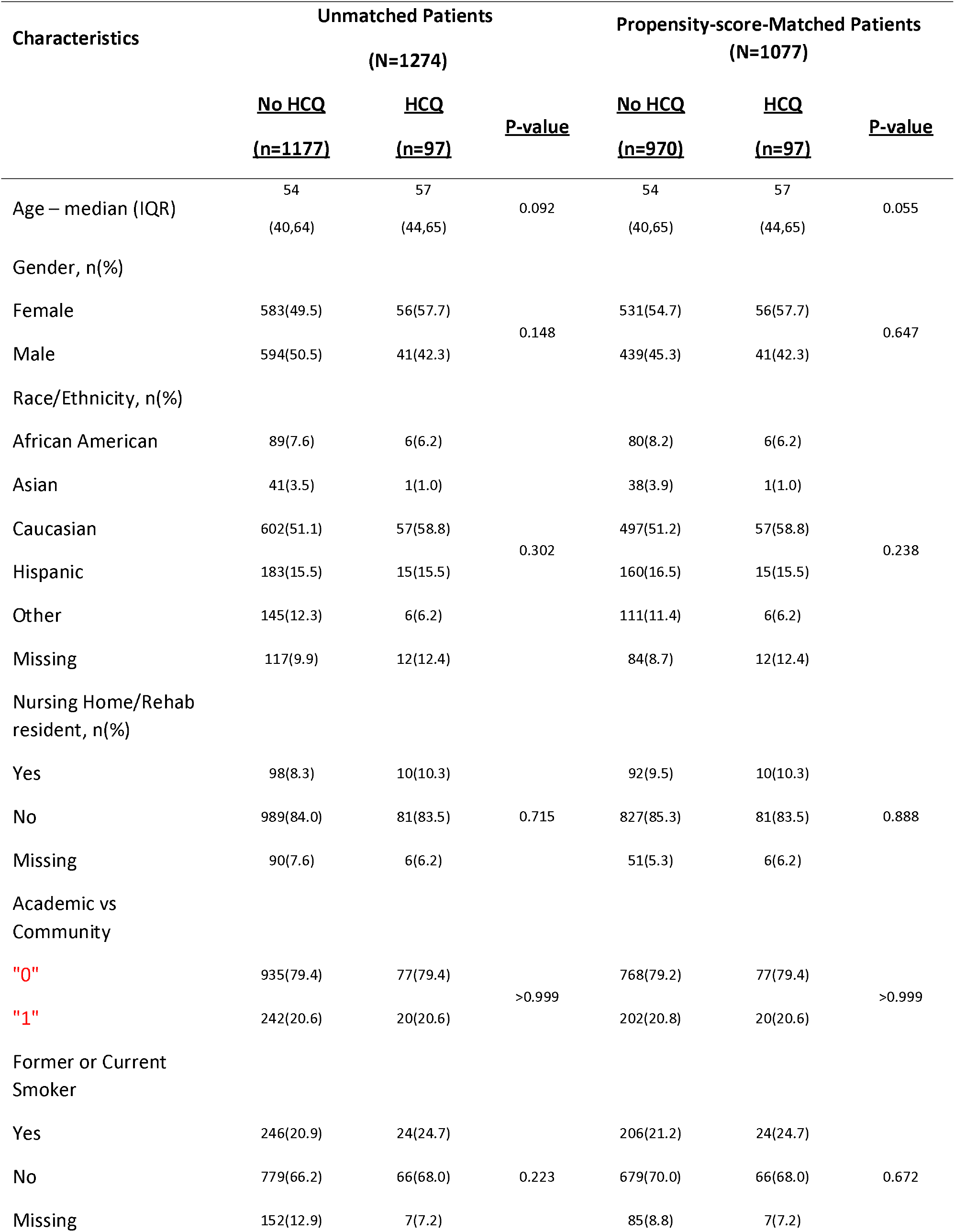
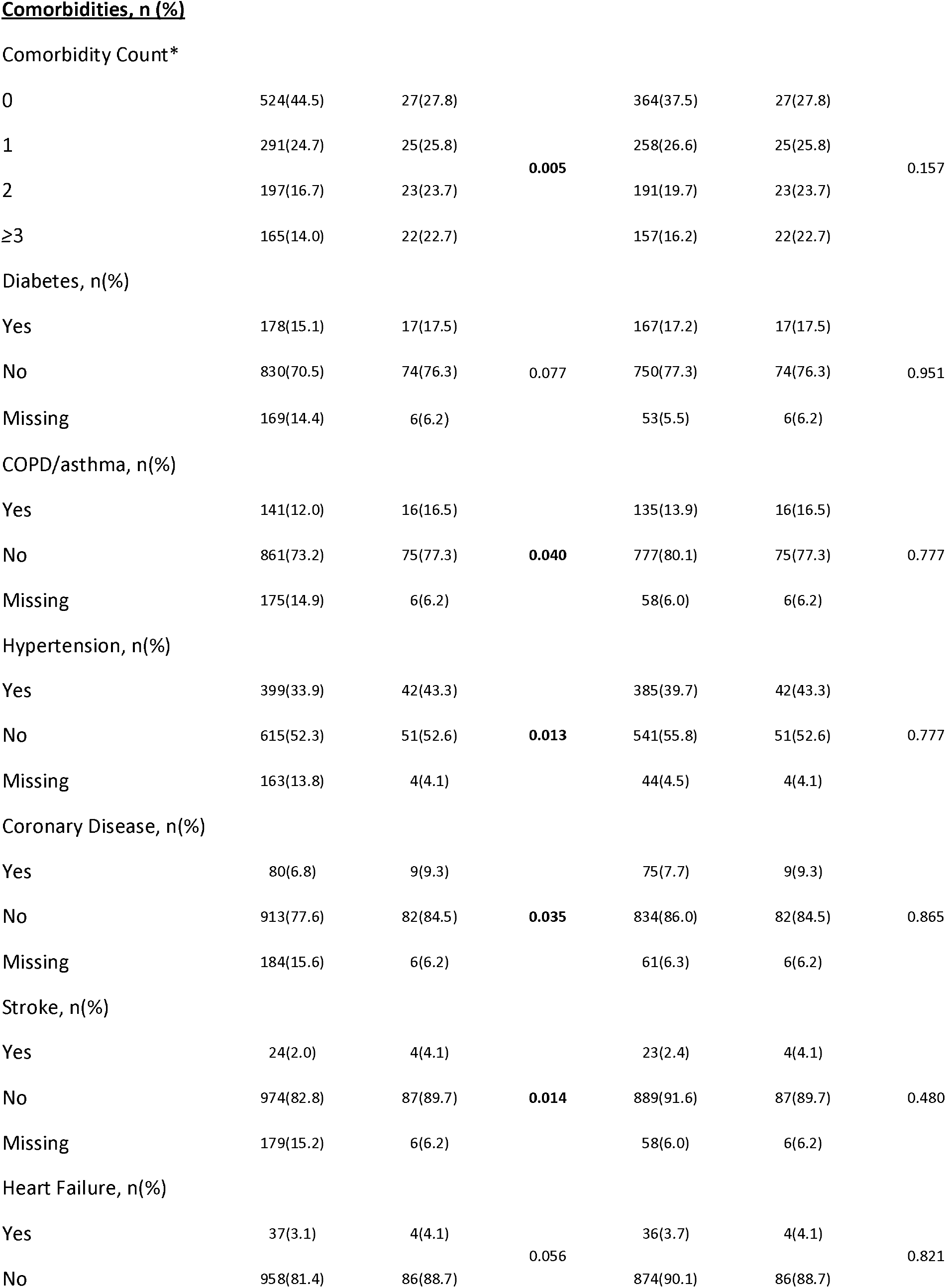
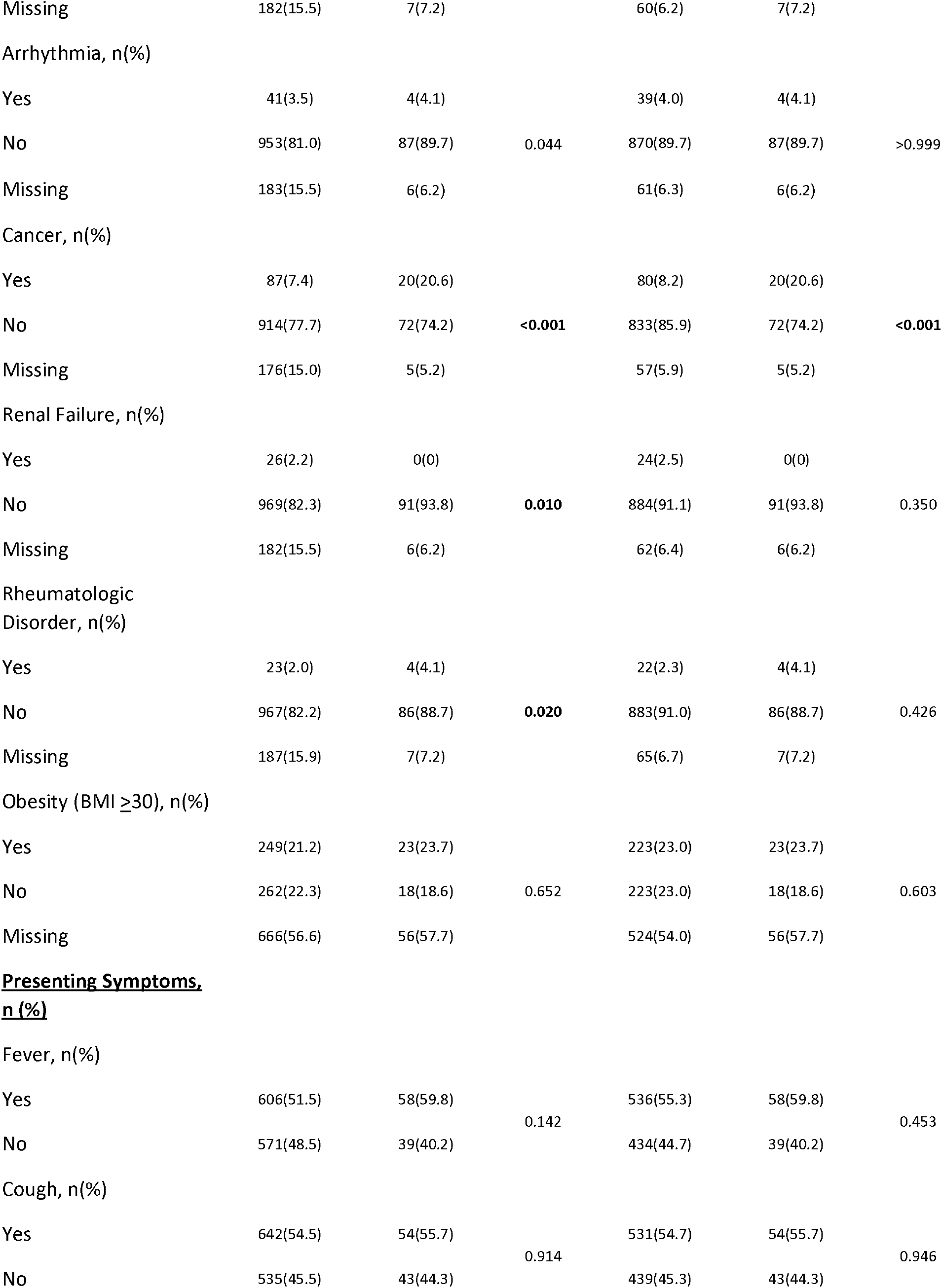
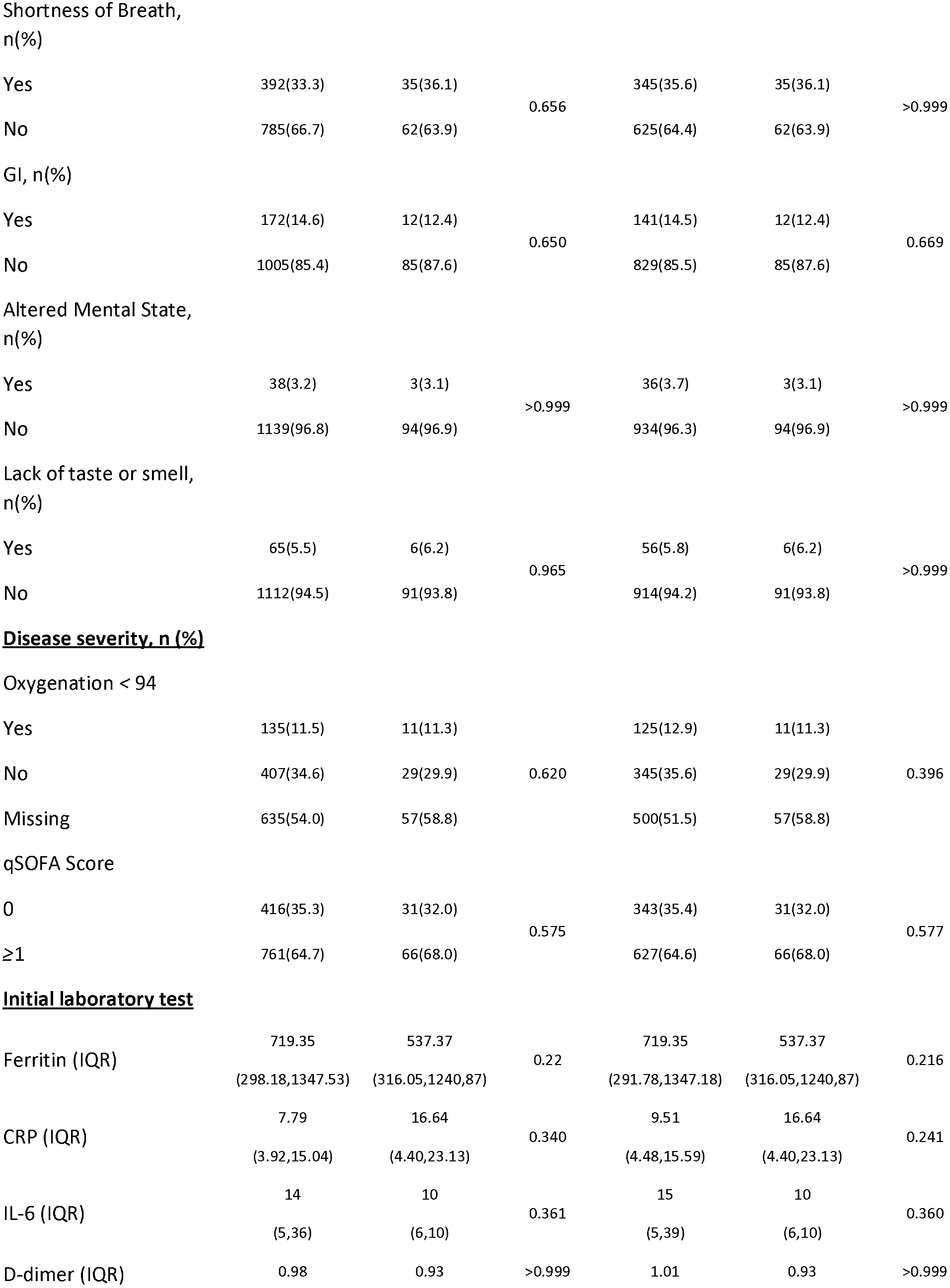
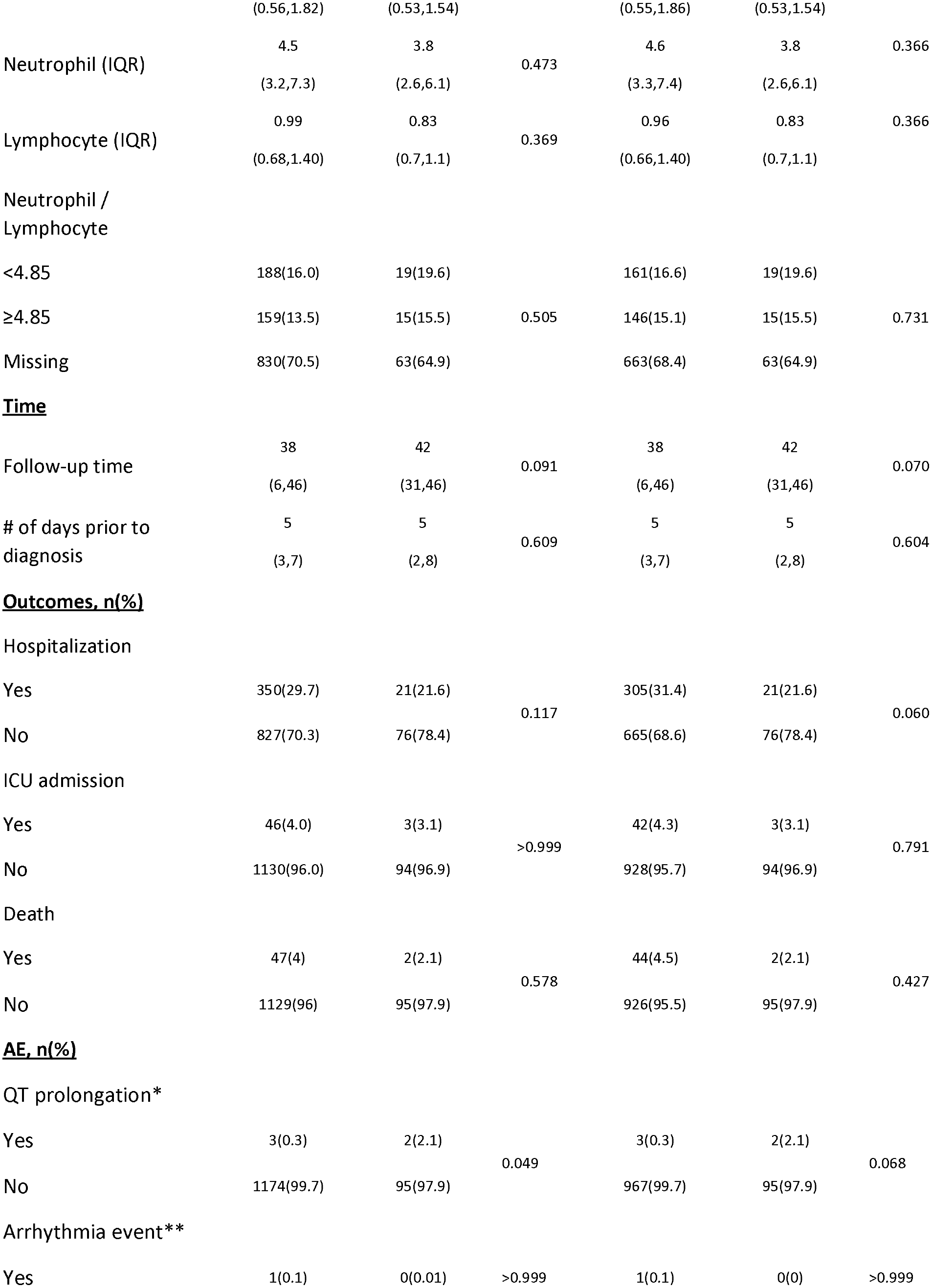
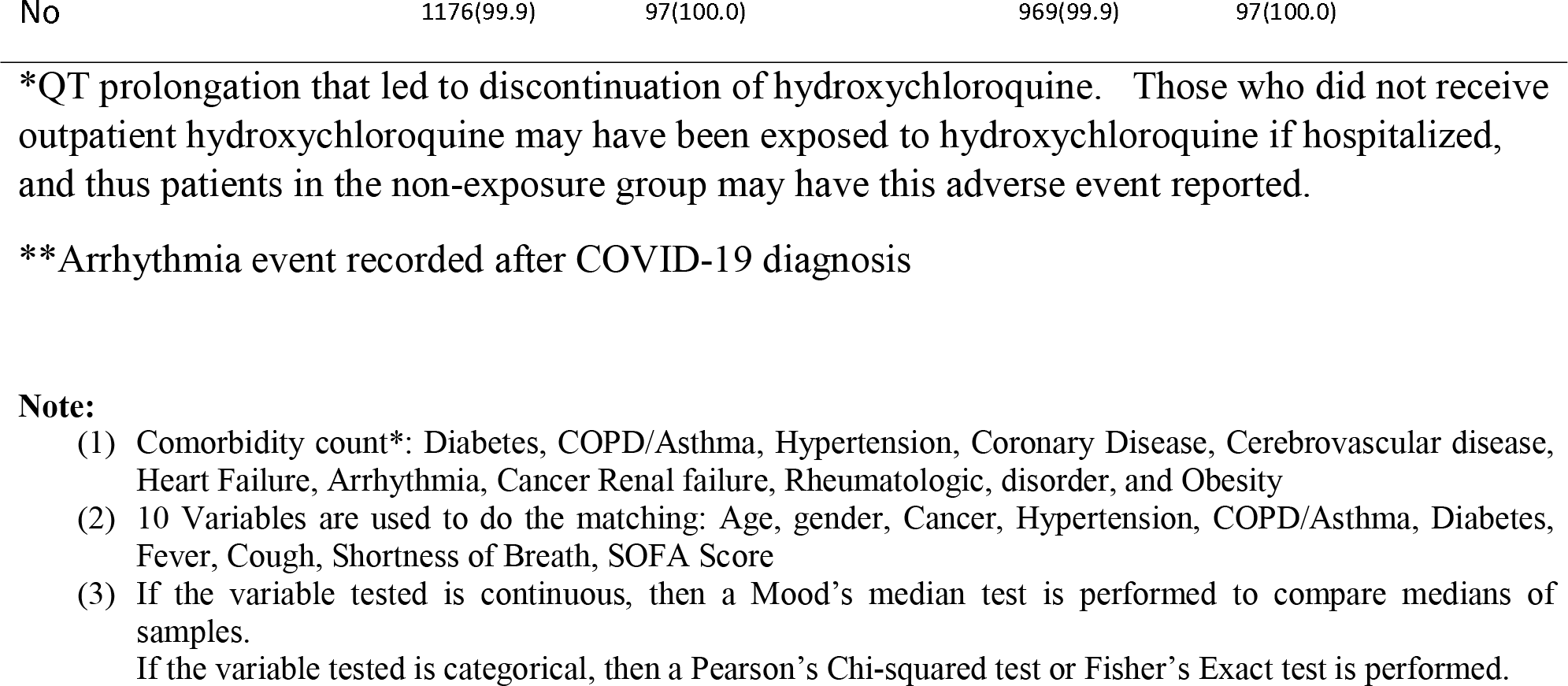
Baseline Characteristics and Outcomes.

In the propensity matched cohort 3 (3.1%) patients with outpatient exposure to hydroxychloroquine subsequently required ICU level support and 42 (4.3%) patients without exposure required ICU care. Ultimately, 2 (2.1%) patients with outpatient exposure to hydroxychloroquine died from COVID-19 related disease and 44 (4.5%) of patients without exposure died (Table 1).

### Primary study endpoints

Among the 1067 outpatients in the propensity matched cohort, with a median of 39 days (IQR 6,46) follow-up, a total of 326 (30.6%) patients required subsequent hospitalization. Three hundred and five (31.4%) patients with no outpatient exposure to hydroxychloroquine were hospitalized and 21 (21.6%) of patients with exposure to hydroxychloroquine were hospitalized. Figure 2 shows the cumulative prevalence of hospitalization from date of diagnosis according to outpatient hydroxychloroquine exposure (log-rank p=0.045). The cumulative prevalence of hospitalization from the self-reported date of onset of symptoms is shown in Supplementary Figure 1 (log-rank p=0.036). 46 (4%) patients with no outpatient exposure required ICU care compared to 3 (3.1%) patients who had outpatient exposure to hydroxychloroquine. 47 (4%) patients with no outpatient exposure died compared to 2 (2%) patients with outpatient exposure to hydroxychloroquine. In patients prescribed hydroxychloroquine as an outpatient for whom follow-up electrocardiographic data were available, QTc prolongation events, defined as discontinuation due to physician discretion, occurred in 2 (2%) of patients, and arrhythmia events after hydroxychloroquine exposure were noted in no patients. (Table 1)

**Figure 2.**
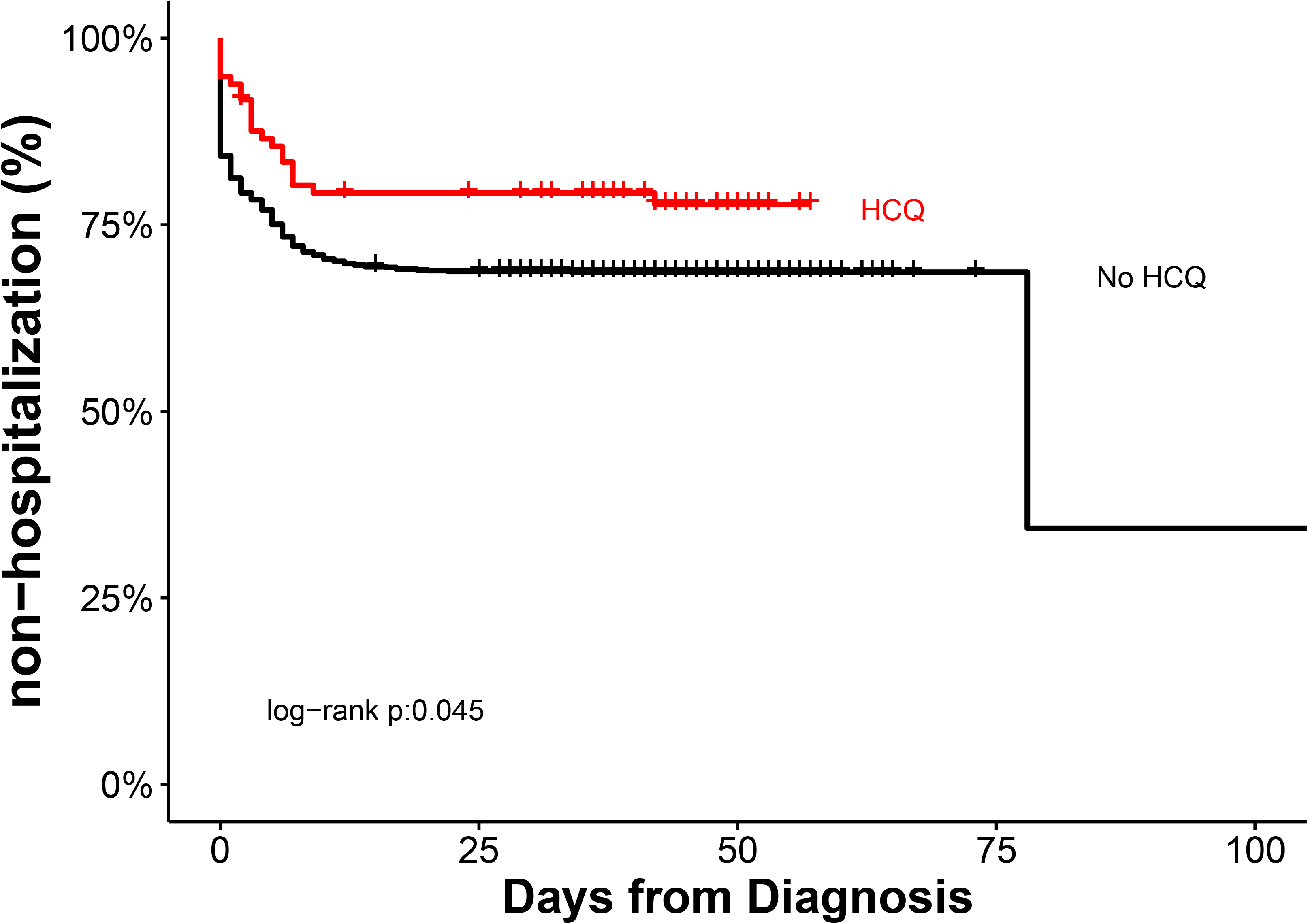
Hospitalization according to Hydroxychloroquine Exposure from Date of Confirmed SARS-CoV-2 Infection. Cumulative prevalence of hospitalization among mildly symptomatic COVID-19 patients according to outpatient exposure to hydroxychloroquine from date of polymerase chain reaction confirmed infection with SARS-CoV-2 in propensity matched cohort HCQ=hydroxychloroquine.

In the primary multivariable logistic regression analysis with propensity matching there was an association between exposure to hydroxychloroquine and a reduced rate of hospitalization related to progressive COVID-19 illness (OR 0.53; 95% CI, 0.29, 0.95) (Table 2). Sensitivity analyses using stepwise (AIC based) variable and Lasso selection yielded similar results in the propensity matched cohorts (Supplementary tables 1-2), and the significant association was also identified in the unmatched cohort (Supplementary tables 3-6).

**Table 2.** Matched multivariable logistic regression model for hospitalization (sample size=1067)

|  | **Estimated**<br>**OR** | **OR 95% CI** | **P**<br>**value** |
| --- | --- | --- | --- |
| HCQ, yes/no | 0.535 | (0.291,0.949) | 0.038 |
| Age | 1.023 | (1.012,1.034) | <0.001 |
| Gender, male/female | 1.316 | (0.957,1.81) | 0.091 |
| Diabetes, yes/no | 1.255 | (0.829,1.895) | 0.281 |
| Hypertension yes/no | 0.941 | (0.644,1.372) | 0.753 |
| COPD/Asthma, yes/no | 0.718 | (0.453,1.126) | 0.154 |
| Cancer, yes/no | 1.045 | (0.607,1.785) | 0.872 |
| **<u>Presenting Symptoms,</u>**<br>**<u>n (%)</u>** |  |  |  |
| Fever, yes/no | 1.265 | (0.866,1.85) | 0.224 |
| Cough, yes/no | 0.853 | (0.573,1.264) | 0.43 |
| Shortness of Breath,<br>yes/no | 6.113 | (4.307,8.765) | <0.001 |
| **<u>Disease severity, n (%)</u>** |  |  |  |
| qSOFA Score, 1/0 | 0.193 | (0.139,0.265) | <0.001 |

Note : Test of model goodness of fit shows a good fit with p-value=0.094 (g=13)

### Exploratory study endpoints

In an exploratory analysis we examined a subgroup of 749 outpatients in the propensity matched cohort who self-reported at least one major symptom of fever, cough or shortness of breath at the time of their time of SARS-CoV-2 diagnosis. In this subgroup 69 (9.2%) patients received hydroxychloroquine prescriptions and 680 (90.8%) patients did not. There were fewer hospitalizations in the hydroxychloroquine cohort (19 patients, 27.5%) compared to individuals with no exposure (259 patients, 38.1%). In the multivariable logistic regression analysis of these symptomatic patients, there was no significant association between hydroxychloroquine exposure and subsequent need for hospitalization (OR 0.74, 95% CI, 0.39, 1.37) (Supplementary table 7, Supplementary figure 2).

Given the strong association between advanced age and subsequent hospitalization requirement in both the unmatched and propensity matched analyses, an additional analysis was conducted on the interaction between age and hydroxychloroquine exposure. Restricting the multivariable logistic regression model to the 282 persons age 65 years or greater resulted in a non-significant odds reduction of hospitalization (OR 0.49, 95% CI 0.17, 1.32). Similar directional trends were seen on sensitivity analyses in this elderly cohort. (Supplementary table 8)

A final subgroup analysis was conducted in patients who were exposed to outpatient hydroxychloroquine according to duration of symptoms, more than 2 days of self-reported symptoms compared to 2 days or less. A univariate logistic regression analysis did not show a significant association with hospitalization, although a directional trend of increased hospitalization was noted when outpatient hydroxychloroquine was administered after more than 2 days of symptoms (OR 3.43, 95% CI 0.57, 66) (Supplementary table 9).

## Discussion

In this multicenter retrospective observational cohort study of mildly symptomatic outpatients with polymerase chain reaction documented SARS-CoV-2 infection, we noted an association (OR 0.53; 95% CI, 0.29, 0.95) between outpatient exposure to hydroxychloroquine and a reduction in subsequent need for hospitalization. Safety events, defined as QT prolongation or arrhythmia occurrence, were minimal, occurring in 2% and 0% of patients. As the majority of COVID-19 patients are mildly symptomatic and treated in outpatient settings, our findings justify further exploration of hydroxychloroquine during this pandemic in this population. If the findings are confirmed, early hydroxychloroquine therapy to a broad outpatient population could have important implications for reducing limited healthcare resources. The economic impact on healthcare might also be significant as the financial cost of a short course of hydroxychloroquine to a large population would be easily recouped by even a modest reduction in hospitalizations.

Our findings in the outpatient setting are in conflict with prior observational studies conducted among hospitalized patients potentially highlighting differences in effect based on the severity of disease.^22^ Following an initial infection by SARS-CoV-2 resulting in attack of alveolar epithelial cells patients may develop a hyper-inflammatory state characterized by activation of the innate immune system and release of pro-inflammatory cytokines and chemokines. Patients who experience this ‘cytokine storm’ progress rapidly to respiratory failure and multi-organ failure.^23-26^ In these hospitalized patients the weak anti-inflammatory effects of hydroxychloroquine may be insufficient to block the cytokine cascade whereas more potent immunosuppressive agents such as dexamethasone and tocilizumab have been associated with beneficial effects.^27-29^

By contrast, hydroxychloroquine has anti-viral effects, decreasing SARS-CoV-2 viral load, and thus may be more suited in preventing the significant tissue damage needed to incite the hyper-inflammatory state.^3,30^ This would position hydroxychloroquine earlier in the clinical course, at the time of early infection, prior to hospitalization need.^31^

As noted above, several recent studies have attempted to explore the role of hydroxychloroquine earlier in the clinical course of COVID-19.^10-14^ However, given enrollment of generally younger patients with low baseline rates of hospitalization, these studies appear under-powered to demonstrate meaningful effects. For example, the recent Spanish randomized trial explored early hydroxychloroquine use, at a median time from symptom onset of 3 days, in the outpatient setting.^10^ While the study did not find a significant decrease in mean viral load up to 7 days after treatment, the investigators reported lower hospitalization rates in the population treated with hydroxychloroquine. Similar non-statistical directional reductions were noted in the other studies. Thus, the potential benefit of hydroxychloroquine in the early management of outpatients should be considered unanswered but of greater interest.

We defined exposure to hydroxychloroquine based on documentation of a prescription being written, but confirmation of prescription fill or full adherence to the complete course was not ascertained, thus mimicking an intention-to-treat model. This limitation biased against finding a difference between cohorts, as non-adherent patients would be categorized within the hydroxychloroquine cohort even though in actuality they did not have drug exposure. Thus, our reduction in hospitalization association may be an underestimate of the effect size. Conversely, it is possible that some outpatients received prescriptions for hydroxychloroquine outside the HMH network and were misclassified in the opposite direction, although this is less likely as patients underwent initial testing within our hospital network and would have been contacted by HMH personnel to discuss testing results and/or had notation of a prescription fill in the EPIC pharmacy section.

Our study was conducted early in the United States pandemic during a timeframe when testing for COVID-19 was largely limited to individuals with symptomatic disease. Thus, we suspect that those included in our observational cohort represent a bias towards more advanced disease with a higher likelihood of hospitalization. Indeed 30.6% of our cohort subsequently required hospital based care, which is higher than current state and national hospitalization rates.^1,2^ Our findings need to be taken into context of current testing availability.

This observational study has several additional limitations. We recorded hospitalizations based on EHR documentation, but we have not accounted for hospitalizations outside the HMH network. Since the patients in our series received outpatient care at an HMH facility we believe that subsequent hospitalizations outside the network were minimal. Observational studies also cannot draw causal inferences given inherent known and unknown confounders. We attempted to adjust for known confounders using our propensity model approach but acknowledge we may not have captured all possible confounders. Misclassifications of the data are possible due to manual abstraction of EHR structured and unstructured data. Missing data, laboratory studies not obtained, and symptoms not reported or documented also limited our analyses. Our study also focused on patients in New Jersey USA, limiting the applicability to other geographic regions with differing treatment and hospitalization algorithms.

In conclusion, hydroxychloroquine exposure among outpatients with mildly symptomatic COVID-19 was associated with a reduction in hospitalization rates from disease progression in this multi-center observational cohort. Further external validation of this finding is required. Although use of hydroxychloroquine in this outpatient population outside the context of a clinical trial cannot be recommended, our study suggests that additional evaluations of hydroxychloroquine are needed in this mildly symptomatic SARS-CoV-2 infected population.

## Data Availability

Deidentified Data may be requested directly from Hackensack Meridian Health. Please contact the corresponding author, Dr. Stuart Goldberg, for specific inquiries.

## Author Contributions

AI, JA, YZ, EH, BAS, UB, SM and SLG had full access to all of the data in the study and take responsibility for the integrity of the data and the accuracy of the data analysis.

Concept and design: AI, SLG

Literature search: AI, AHG, ALP, SLG

Figures: AI, JA, YZ, SLG

Study Design: AI, JA, YZ, AHG, ALP, ISS, SLG

Data Collection and Analysis: all authors

Data Interpretation: AI, JA, YZ, EH, SM, SLG

Writing: all authors

## Acknowledgements

We would like to personally thank all of the data abstracters, nurses, and physicians who helped collect data for this study. This study was not funded by any external sources. None of the authors are employed by the National Institute of Health or are in receipt of any National Institute of Health grants having to do with this manuscript.

